# Pathogenwatch: A public health platform for rapid interpretation of pathogen genomics

**DOI:** 10.64898/2026.03.18.26348693

**Authors:** Nabil-Fareed Alikhan, Corin Yeats, Khalil Abudahab, Pranit Shinde, Georgina Lewis-Woodhouse, Anthony Underwood, Silvia Argimón, Ravikumar K Lingegowda, Pilar Donado-Godoy, Sonia Sia, Iruka N Okeke, Sophia David, Philip M Ashton, David M Aanensen

**Affiliations:** Centre for Genomic Pathogen Surveillance, Pandemic Sciences Institute, University of Oxford, Oxford, UK; WHO Collaborating Centre on Genomic Surveillance of AMR, University of Oxford, Oxford, UK; KIMS Hospital and Research Centre, Bangalore, India; Global Health Research Unit for the Genomic Surveillance of Antimicrobial Resistance, CI Tibaitatá, Corporación Colombiana de Investigación Agropecuaria (AGROSAVIA), Mosquera, Colombia; Research Institute for Tropical Medicine, Manila, Philippines; Department of Pharmaceutical Microbiology and Biotechnology, University of Ibadan, Ibadan, Nigeria; NIHR Global Health Research Unit on Genomics and enabling data for surveillance of antimicrobial resistance

## Abstract

Pathogen genomic data provide important insights for public health microbiology, yet genome analysis options often remain highly technical and beyond the reach of many microbiologists and public health practitioners. Pathogenwatch (https://pathogen.watch) is a platform that translates pathogen genome data into outputs directly usable for surveillance and public health action. The platform contextualises bacterial, viral, and fungal genomes within a unified framework integrating organism identity, variant or lineage assignment, antimicrobial resistance and virulence gene detection, and geographic and temporal context. Pathogenwatch provides multilocus sequence typing (MLST) for more than 37 bacterial species and core genome MLST (cgMLST) schemes for over 20 priority organisms, with user-uploaded genomes automatically compared against over 875,000 curated public bacterial genomes. The platform has been adopted by 14,389 registered users across 165 countries. In 2025, users uploaded 328,676 genome assemblies and 20,830 read datasets. Pathogenwatch replicates analysis results of complex bioinformatics pipelines. Benchmarking of SARS-CoV-2 lineage assignment against an established reference dataset demonstrated complete concordance for all Variants of Concern and Interest, and full concordance with contemporary Pangolin calls across non-VOC/VOI lineages. Pathogenwatch operates as a continuously deployed, containerised system designed for scalability, reproducibility, and rapid incorporation of new pathogens, positioning it as durable infrastructure for both endemic surveillance and genomic response to emerging threats.

## Introduction

Whole-genome sequencing has transformed the study of infectious diseases by enabling high-resolution reconstruction of pathogen evolution, transmission, and population structure (1). Comparative analyses of large genomic datasets now support fine-scale discrimination of transmission chains, identification of emerging variants and lineages, and tracking of genomic markers relevant to public health across spatial and temporal scales (2). As sequencing capacity has expanded globally, 2.4 million of pathogen genomes have been deposited in public archives such as the NCBI Sequence Read Archive (SRA), creating unprecedented opportunities for genomic epidemiology and public health surveillance (1,3–5).

Despite the increasing use of sequencing data, the translation of genomic data into routine public health action remains uneven (1). Many genome sequences are generated without access to analytical frameworks that allow users to relate genetic similarity to geography, time, and inferred phenotypic traits such as antimicrobial resistance or virulence.

Pathogenwatch (https://pathogen.watch) has been iteratively developed through collaborations with community experts for specific species, including *Salmonella enterica* serovar Typhi (6), *Klebsiella* (7) *Neisseria gonorrhoeae* (8) and *Streptococcus pneumoniae* (9), with the aim of creating generic pipelines that can be applied easily to other species. The focus on re-use and modularity means Pathogenwatch supports the identification and assignment of MLST types to over a hundred species, along with detailed genotyping of more than a dozen key species of concern.

Here, we describe Pathogenwatch as a continuously operating, general-purpose genomic surveillance platform designed to support public health genomics across bacterial, viral, and fungal pathogens. Central to this transition is the adoption of a common public health abstraction in which genome sequences are transformed into interpretable signals defined by organism identity, variant or lineage assignment, and relevant genomic markers integrated with geographic and temporal context. This abstraction enables a consistent user interface to be deployed across biological kingdoms while supporting species-appropriate analytical resolution. This design explicitly addresses the need for surveillance systems that can rapidly pivot to novel pathogens during public health emergencies, without requiring re-engineering of core analytical infrastructure.

Furthermore, Pathogenwatch enables user-uploaded genomes to be contextualised against a large and continuously maintained collection of public reference data. Genomes submitted by users are analysed on demand and placed within pre-computed, quality-controlled global datasets that are automatically assembled from primary sequence repositories. These public datasets are routinely filtered to retain high-quality genomes with the minimal metadata required for meaningful epidemiological interpretation. Integrated visualisation of phylogenetic relationships, spatial distribution, temporal trends, and genomic features allows users to explore and compare their data within a global context, without requiring local bioinformatics infrastructure. These capabilities are underpinned by a scalable, containerised architecture designed to support reproducibility, extensibility, and community-driven development.

## Methods

### Platform architecture

Pathogenwatch is a cloud-native, containerised platform engineered for scalability, reproducibility, and multi-organism support, with its primary deployment implemented on Amazon Web Services (AWS). Analytical workflows are packaged as containers stored in AWS Elastic Container Registry (ECR) and orchestrated using AWS-managed container services (including ECS with Fargate), enabling seamless execution across cloud, cluster, and local environments. This architecture ensures consistent execution of pipelines while supporting continuous ingestion of public datasets and elastic scaling of compute resources during periods of peak demand.

### Speciation with Speciator

Speciator is Pathogenwatch’s in-house species assignment module, providing automated classification of assembled genomes to support downstream species-specific analyses such as cgMLST and antimicrobial-resistance prediction. It uses a MinHash-based k-mer search approach (via Mash (10)) combined with curated reference genome libraries drawn from established resources and in-house curation, enabling rapid and reliable species identification across a wide range of bacterial taxa.

Species assignment is performed using a multi-stage reference-based strategy designed to balance speed, accuracy, and robustness, with fallback behaviour for novel or poorly represented taxa. Benchmarking across large curated genome collections demonstrated high classification accuracy and execution times of only a few seconds per assembly, supporting scalable, automated surveillance workflows. Full methodological details, including reference library construction, distance thresholds, and fallback logic, are provided in the Supplementary Methods.

### Genome Assembly

Paired-end Illumina reads are assembled de novo using a Nextflow-implemented SPAdes pipeline that supports execution on local, cluster, or cloud environments (11). Submissions are handled through the Pathogenwatch runner system, with fair-use scheduling to manage computational load. The workflow includes quality control (FastQC (12)), trimming (Trimmomatic (13)), error correction (Lighter (14)), contamination screening (ConFindr (15)), read merging (FLASH (16)), and SPAdes assembly (17).

### Genome quality control in Pathogenwatch

Genome quality control in Pathogenwatch is performed using species-specific thresholds derived from the QualiBact framework (18). QualiBact defines reproducible quality criteria for bacterial genome assemblies based on empirical distributions of standard assembly and completeness metrics observed across large public datasets. Quality assessment is based on a combination of assembly statistics and completeness estimates, including total assembly length, N50, number of contigs, GC content, and estimates of completeness and contamination. Species-specific acceptable ranges for these metrics were derived in QualiBact using large-scale public genome collections and curated reference genomes. Within Pathogenwatch, assemblies that fall outside species-appropriate quality ranges are flagged during quality control.

### Contextual clustering and lineage assignment

Pathogenwatch infers genomic relatedness and lineage structure through a combination of core-genome multilocus sequence typing (cgMLST), hierarchical clustering (HierCC) (19) for *Escherichia coli* and *Salmonella*, LIN codes (20), and phylogenetic visualisation tools.

Core-genome MLST (cgMLST) is the primary genotyping framework, implemented using species-specific schemes (typically comprising 1,500–3,000 loci). Each genome is assigned an allelic profile across the defined core set, allowing rapid comparison and clustering of isolates within and across species. CgMLST-based searches and clustering are computed dynamically within individual user contexts rather than relying on a single, fixed global clustering, allowing flexible interpretation as reference datasets and collections evolve.

Related genomes are identified using cgMLST allele distances through the hclink module, which performs dynamic single-linkage clustering. In a context search with hclink, a query genome is first compared against all genomes within the selected public or shared folders using a user-defined allele distance threshold. The closest matching genomes (up to 5,000) are then clustered using single-linkage clustering. Context searches are accessible from both single-genome reports and cluster views. The single-genome report provides a preview of related clusters and links to an interactive viewer with integrated metadata and annotation, while in the collection view, related genomes can be added to a new or existing collection for downstream analysis.

### Additional analytics

Pathogenwatch incorporates a comprehensive suite of post-assembly analytics for the identification of genomic features with public health or clinical relevance, including serotypes, antimicrobial resistance determinants, virulence-associated genes, and lineage-defining markers. Plasmid content is annotated using Inctyper (21) and PlasmidFinder (22), which identify plasmid replicon types and incompatibility groups, supporting the investigation of mobile genetic elements and the dissemination of antimicrobial resistance.

Serotyping and lineage assignment are implemented through species-specific genotyping schemes, including ECtyper (23), SISTR (24), Genotyphi (25), ClermonTyper (26), and Kaptive (27), depending on the organism analysed. These tools predict classical antigenic types—such as O and H antigens in *Salmonella* or K and O loci in *Klebsiella*—as well as multilocus sequence types (MLST or cgMLST), enabling consistent definition of lineages and population structure.

Antimicrobial resistance (AMR) detection combines curated rule-based methods with database-driven approaches using AMRFinderPlus (28), Kleborate (29), and the in-house Pathogenwatch-AMR workflow (30). These analyses identify acquired resistance genes, resistance-associated chromosomal mutations, and plasmid-borne determinants, producing drug-class-level resistance summaries informed by plasmid annotations and aligned to clinical relevance.

Virulence profiling is conducted using modules including VirulenceFinder (31), Vista virulence (32), and species-specific virulence marker sets. These analyses identify key virulence-associated loci and provide contextual information to support surveillance, outbreak investigation, and risk assessment. Collectively, these integrated analytics enable comprehensive and reproducible genome interpretation—from lineage assignment and population structure to resistance and virulence gene content—within a unified Pathogenwatch framework.

### Staphylococcus aureus comparison

*Staphylococcus aureus* ST239 genome assemblies were analysed using Pathogenwatch (https://pathogen.watch). Genomes were processed with the species-specific workflow, including quality control, core-genome phylogenetic reconstruction, and antimicrobial-resistance prediction using Pathogenwatch AMR (30), with results visualised alongside geographic metadata.

### SARS-CoV-2 comparison

SARS-CoV-2 benchmark datasets were obtained from the standardised lineage datasets described by Xiaoli *et al.* (33), including both VOC/VOI and non-VOC/VOI representative genomes. Genome assemblies and associated metadata were analysed using Pathogenwatch’s SARS-CoV-2 workflow. Lineage assignments generated by Pathogenwatch were compared against the lineage labels provided in the benchmark dataset metadata and against Pangolin lineage calls. Strain names were normalised prior to comparison to ensure consistent matching across datasets. Concordance was assessed on a per-genome basis.

## Results

### Pathogenwatch as a general-purpose genomic epidemiology platform

Pathogenwatch is an organism-agnostic genomic surveillance platform that supports bacterial, viral, and fungal pathogens within a consistent analytical framework. The platform operates in near real-time, continuously contextualising newly analysed genomes against global reference datasets and enabling interpretation across diverse pathogens using shared principles (Figure 1).

**Figure 1:**
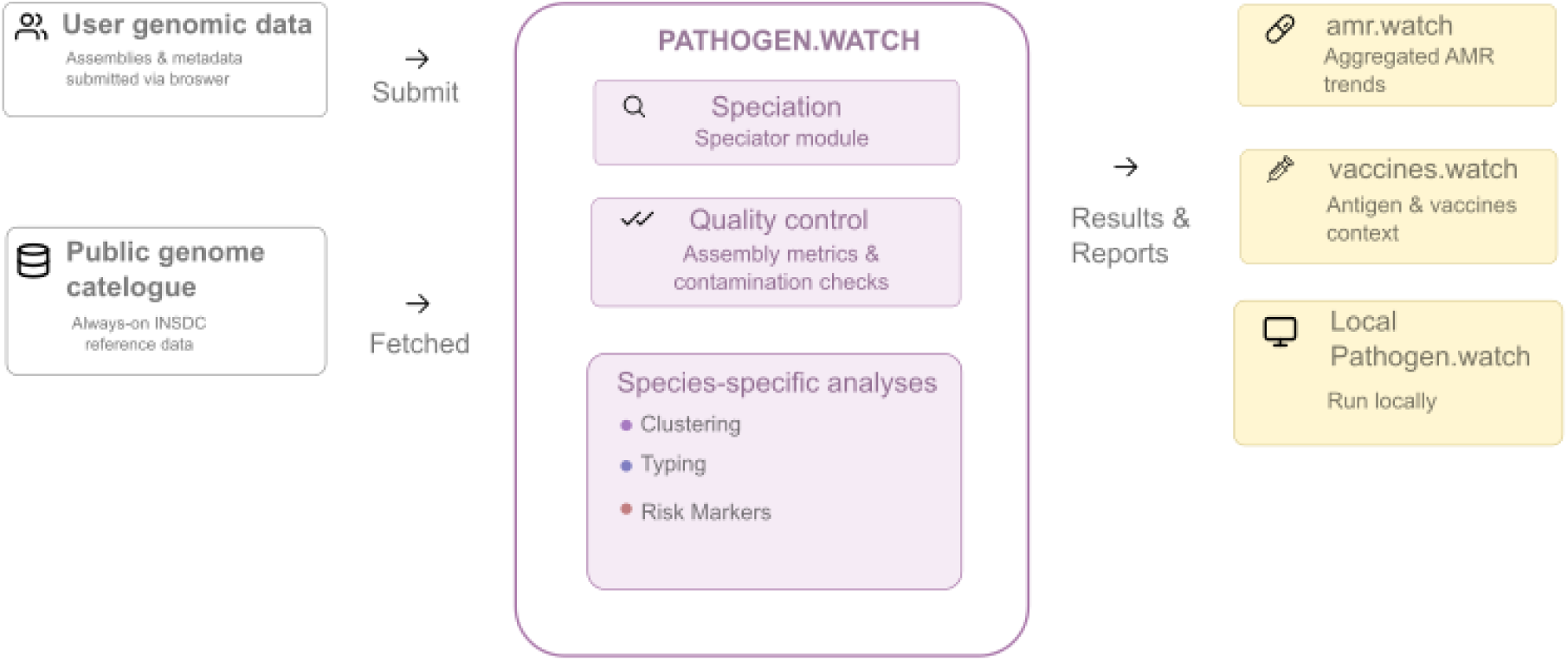
Pathogenwatch working environment. User-submitted genomic data and metadata are processed alongside continuously ingested public genomes from international sequence archives (INSDC: ENA/EBI, NCBI, DDBJ). Incoming genomes first undergo automated speciation and quality control before being analysed using species-specific pipelines. These analyses include contextual searching against curated global population frameworks, inference of evolutionary relationships, genotype or serotype assignment, and antimicrobial resistance (AMR) prediction. Results from public genomics (from INSDC) are integrated into a genomic catalogue that provides up-to-date global context for interpretation. These aggregated outputs are shared with downstream resources, including amr.watch for AMR trend analysis and vaccines.watch for antigenic and vaccine context, while the same workflows can be run locally via Pathogenwatch Local. This modular, containerised architecture supports scalable, reproducible analyses across bacterial, viral, and fungal pathogens and enables elastic scaling during periods of increased demand.

Central to this approach is the separation of core public health questions from organism-specific biology. For all pathogens, Pathogenwatch derives a common set of signals which include organism identity, lineage or variant assignment, and relevant genomic markers. These are interpreted in their geographic and temporal context. Species-specific analyses are applied only where required, allowing Pathogenwatch to support heterogeneous pathogens without fragmenting analytical standards.

Pathogenwatch has been widely adopted as shared infrastructure for genomic surveillance, with 14,389 registered users and 11,013 having uploaded genomic data as of January 2026. In 2025, users uploaded 328,676 genome assemblies and 20,830 read datasets, supporting both routine surveillance and outbreak-driven analyses. Platform usage spans 165 countries, with sustained activity across all inhabited continents and an average of approximately 147 unique users per day, demonstrating continuous global engagement.

In Pathogenwatch, major bacterial pathogens of public health importance, including *Escherichia coli*, *Salmonella enterica*, *Listeria monocytogenes*, and WHO priority pathogens (34) benefit from expanded and harmonised analytical pipelines that extend beyond earlier Pathogenwatch implementations. These include improved lineage frameworks, broader detection of genomic risk markers, and refined antimicrobial-resistance prediction (Table 1).

**Table 1:**
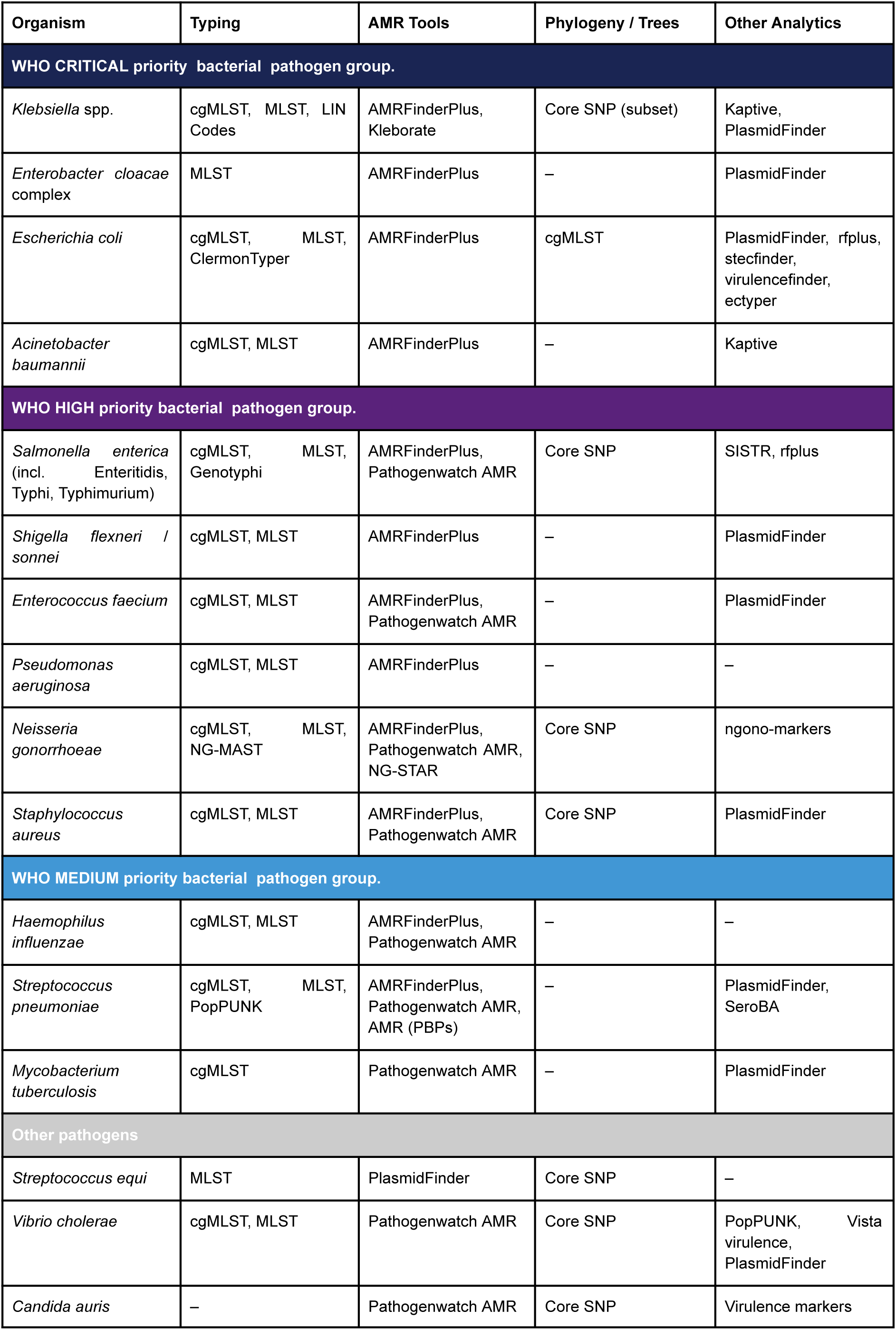
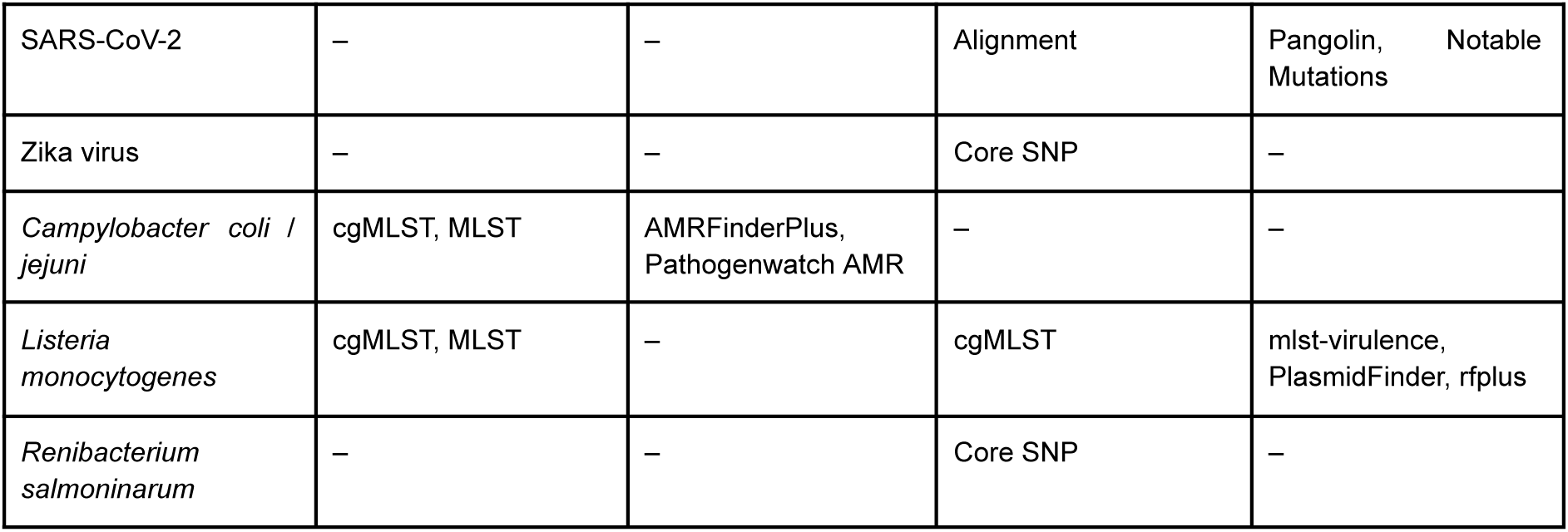
Enhanced organism schemes in Pathogenwatch.

Recent integration of QualiBact-derived (18) thresholds enables automated, species-aware quality control across expanding genomic datasets. Marked interspecies differences in genome size, GC content, and assembly contiguity highlight the need for species-specific criteria rather than uniform cutoffs. Applying these thresholds consistently to both user-submitted and public genomes ensures biologically comparable data for downstream contextual and population-level analyses.

### Scalable infrastructure enabling real-time genomic surveillance

Scalable infrastructure is essential to support real-time genomic surveillance at global scale. Pathogenwatch is designed to operate as a continuously deployed system in which genomic data ingestion, analysis, and contextualisation occur as an integrated, automated process, enabling timely interpretation of new data without manual intervention. Recent internal improvements (see Methods) allow Pathogenwatch to scale computational resources dynamically in response to demand and to deploy analytical software using version-controlled containers, ensuring reproducibility, operational consistency, and alignment with community-standard workflows.

Pathogenwatch contextualises newly uploaded genomes through hclink (see Methods), which identifies related public and private genomes using cgMLST-based context searching against curated, species-specific population frameworks. Unlike fixed global clustering approaches, hclink performs dynamic single-linkage clustering, allowing genomes to be interpreted flexibly against evolving reference datasets. This provides immediate context for interpreting lineage membership, geographic spread, and associated resistance or virulence profiles.

Contextual searching is enhanced by the continuous ingestion and processing of publicly available genomic data from INSDC archives (EBI, NCBI, DDBJ). Genomes retrieved from international sequence archives are analysed using the same standardised pipelines applied to user-submitted data, ensuring direct comparability. Over 1,759,554 bacterial genomes are now included. These data are also delivered through amr.watch (35) and vaccines.watch (36) platforms, which provide complementary, population-level views of genomic surveillance data. Continuous expansion of these reference datasets, alongside improvements in search performance and scalability, enables users to contextualise new data against an increasingly comprehensive global backdrop. These developments support routine surveillance use cases as well as rapid exploratory analyses during outbreaks or emerging events.

Yet, consistent data processing with provenance is vital to keep this volume of data comparable. To demonstrate how Pathogenwatch’s containerised, continuously deployed workflows enable reproducible and standardised genomic analysis at scale, we evaluated SARS-CoV-2 lineage assignment in Pathogenwatch using an established benchmark dataset. This dataset included SARS-CoV-2 genomes of both Variant of Concern/Interest (VOC/VOI) and non-VOC/VOI lineages (33).

Across the VOC/VOI dataset (16 genomes), Pathogenwatch showed complete concordance with the benchmark annotations and Pangolin lineage calls. All major epidemiologically significant lineages, including Alpha (B.1.1.7), Beta (B.1.351), Gamma (P.1), Delta (B.1.617.2), and related VOI lineages, were correctly identified. This demonstrates reliable performance for the detection and classification of high-priority variants relevant to public health surveillance.

In the non-VOC/VOI dataset (39 genomes), Pathogenwatch lineage assignments were fully concordant with Pangolin across all analysed genomes (37). In several cases, discrepancies were observed between Pathogenwatch (which used Pangolin v4.3) and the original dataset annotations (33), reflecting updates and refinements in lineage definitions since the creation of the benchmark datasets, which used Pangolin v3.1.3. In these instances, Pathogenwatch assignments matched contemporary Pangolin calls (37), indicating alignment with current nomenclature.

Overall, these results demonstrate that Pathogenwatch supports routine SARS-CoV-2 surveillance through its continuously operating deployment, enabling lineage assignments to be interpreted in the context of an up-to-date, globally contextualised genomic framework.

### User-facing analytical evolution and enhanced comparative analysis

The evolution of Pathogenwatch into a generic, multi-organism platform has been accompanied by targeted improvements to user-facing analytical capabilities, designed to support comparative genomic epidemiology across diverse pathogens while remaining accessible to users with varying levels of bioinformatics expertise (Figure 2).

**Figure 2:**
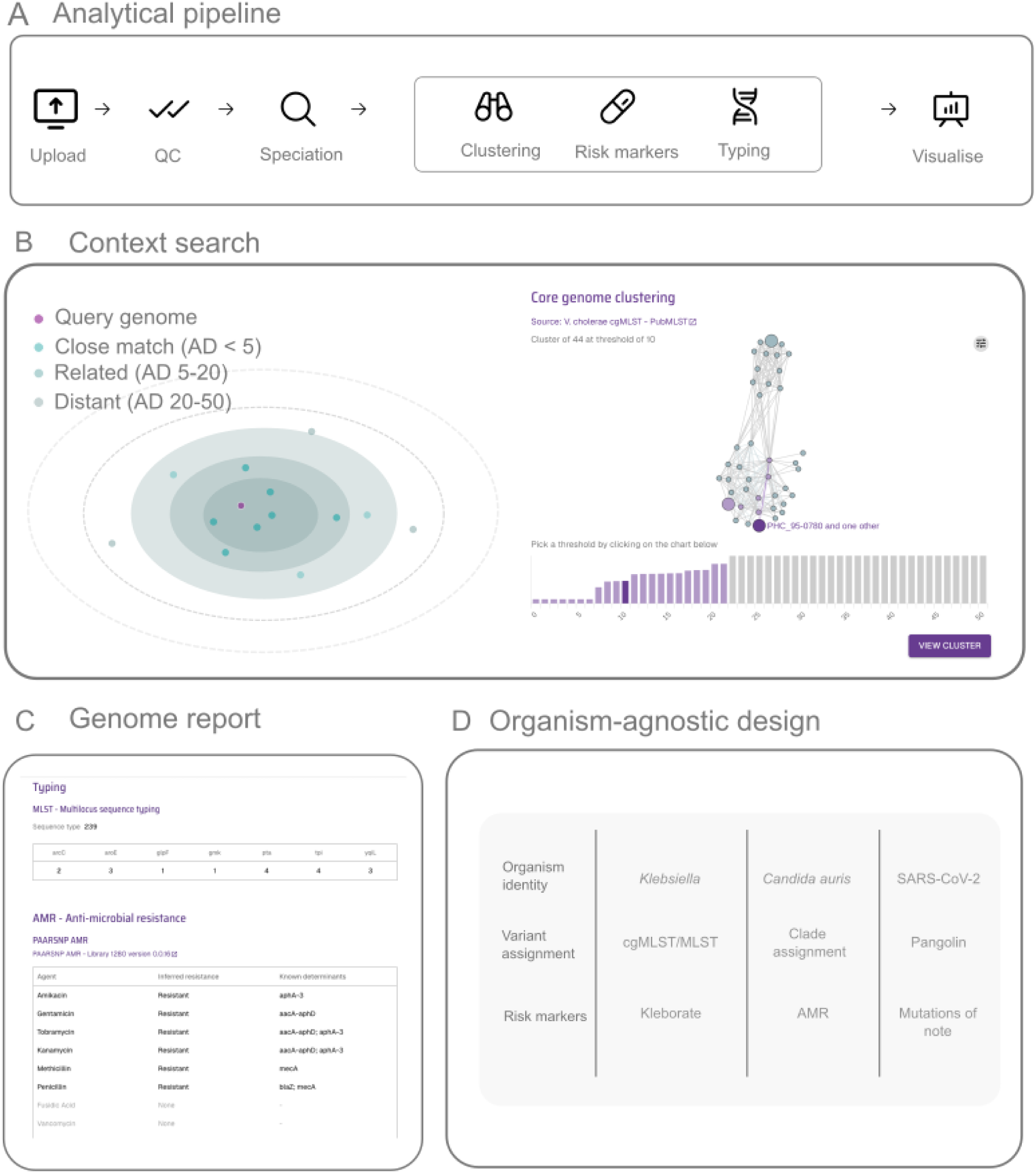
Pathogenwatch analytical views and user workflows. (a) Genomic data enters a modular analytical pipeline encompassing quality control, speciation, clustering, risk marker detection and genotyping against curated public datasets. (b) Context Search uses the hclink module to perform cgMLST-based distance (AD: Allele distance), using single lineage clustering. Matched isolates are displayed within a population framework alongside their geographic distribution, enabling rapid identification of closely related lineages, resistance profiles, and geographic spread. (c) Individual Genome Reports summarise quality metrics, genotype and sequence type assignments, AMR predictions, and plasmid/virulence annotations for a single genome. (d) Pathogenwatch applies a consistent analytical interface across bacterial, viral, and fungal pathogens while deploying species-appropriate typing schemes, AMR detection tools, and genotyping frameworks. Representative organisms shown: *Klebsiella*, SARS-CoV-2, and *Candida auris*.

Pathogenwatch presents genomic epidemiological analyses through a set of dynamically linked views that integrate genetic relatedness, geographic distribution, temporal context, and inferred genomic features. Individual “Genome Reports” summarise lineage assignment, quality metrics, and resistance predictions, while “Collections” provide the primary environment for comparative analysis across outbreaks, surveillance cohorts, or research datasets. Within Collections, phylogenetic trees, maps, timelines, and metadata tables are interactively linked, allowing users to explore how genetic relatedness corresponds to patterns of spread, persistence, and genomic risk factors.

Recent enhancements to the Collection View improve clarity and flexibility across organisms, including direct visualisation of metadata on trees and other views. These features enable rapid identification of clusters, outliers, and emerging variants, and support interpretation in mixed-organism or multi-context surveillance settings. Public reference genomes can be incorporated seamlessly to provide immediate global context, while user-submitted data remain private unless explicitly shared.

To demonstrate these capabilities, we reanalysed a well-characterised collection of *Staphylococcus aureus* genomes originally described by Harris *et al.* (38), which examined the global dissemination and hospital transmission of the multidrug-resistant MRSA lineage ST239 (Figure 3). Core-genome SNP phylogenetic reconstruction reproduced the strong geographic structure reported in the original study, with isolates clustering largely by country and continent, consistent with limited intercontinental transmission followed by regional clonal expansion.

**Figure 3:**
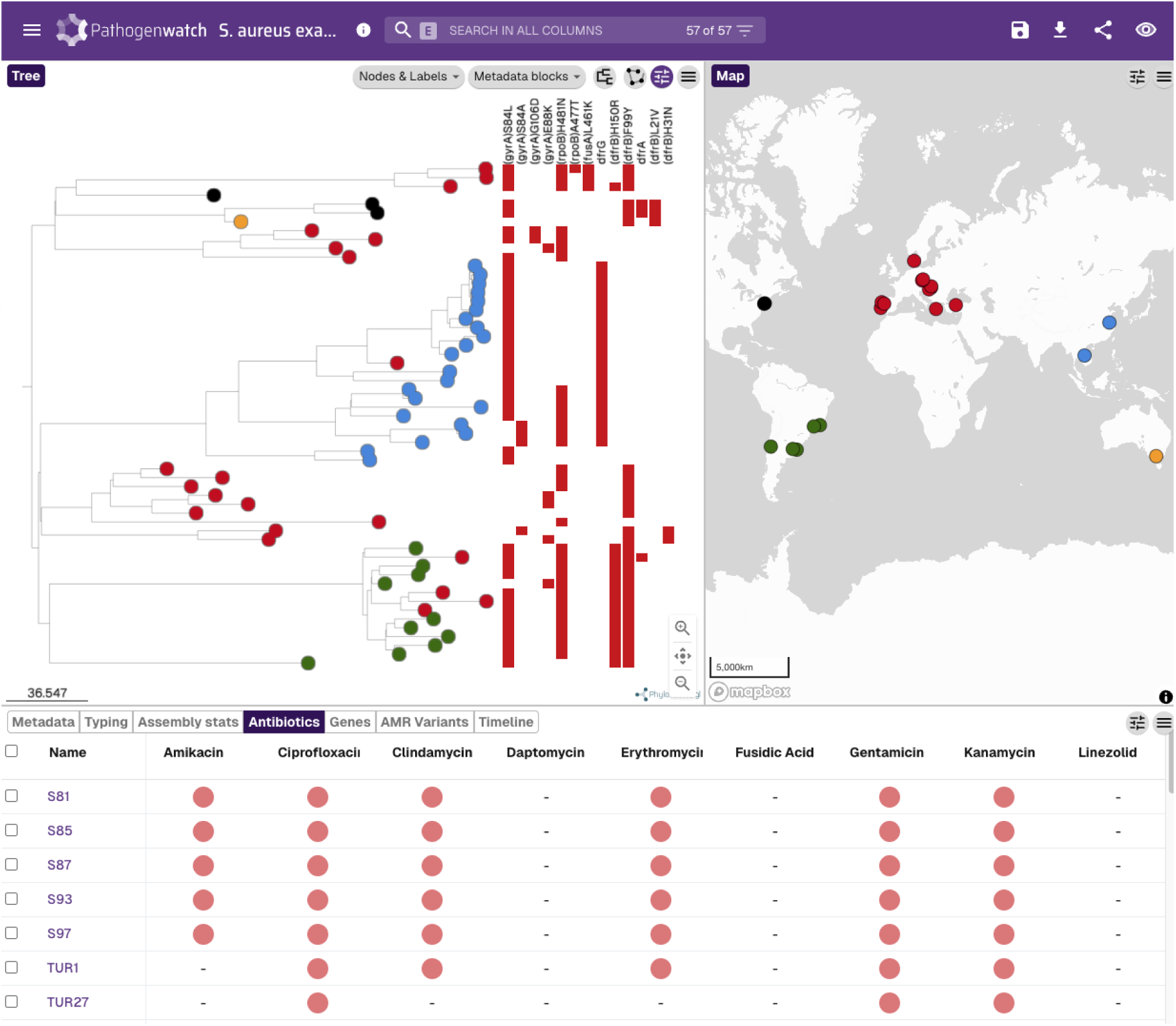
Pathogenwatch analysis of a global *Staphylococcus aureus* ST239 dataset. Screenshot of the Pathogenwatch Collection View showing integrated phylogenetic, geographic, and antimicrobial-resistance analyses. The core-genome phylogeny (left) reveals continent- and country-level clustering of isolates, consistent with global population structure, while the map (right) shows their geographic distribution. The lower panel summarises predicted antimicrobial resistance profiles inferred using Pathogenwatch AMR workflows. Together, these linked views illustrate how Pathogenwatch enables contextual interpretation of genomic relatedness, spatial patterns, and resistance determinants within a single surveillance interface. The collection is available at https://next.pathogen.watch/collections/86HqKQHQSkqLrEyPAzB3Yv-s-aureus-example

Pathogenwatch also identified clinically relevant antimicrobial-resistance variation within the ST239 lineage. Resistance-associated chromosomal mutations in loci including *gyrA*, *grlA*, *rpoB*, *fusA*, and *dfrB* were detected using the Pathogenwatch AMR workflows, with their distribution across the phylogeny reflecting repeated, independent emergence under antimicrobial selection pressure.

## Discussion

The results presented here demonstrate that Pathogenwatch has matured into a continuously operating, organism-agnostic surveillance platform with broad global adoption: over 14,000 registered users across 165 countries collectively uploaded more than 300,000 genome assemblies in 2025 alone, and the platform now contextualises user data against over 1.7 million curated public bacterial genomes. Benchmarking of SARS-CoV-2 lineage assignment confirmed complete concordance with established standards, while reanalysis of a global Staphylococcus aureus ST239 collection reproduced known phylogeographic structure and identified clinically relevant resistance variation within the lineage. Together, these findings validate Pathogenwatch as durable, system-level infrastructure for genomic epidemiology—capable of supporting bacterial, viral, and fungal pathogens within a unified analytical framework while lowering barriers to routine genomic surveillance.

A key contribution of the platform is the adoption of a common public health abstraction through which genome sequences are transformed into interpretable signals with organism identity, lineage or variant assignment, and relevant genomic markers integrated with user-submitted geographic and temporal context. This approach enables consistent interpretation across diverse pathogens while retaining species-appropriate analytical resolution. Continuous ingestion of public genomic data and dynamic contextualisation allow new genomes to be interpreted immediately against an up-to-date global reference, supporting both routine surveillance and outbreak investigation.

The containerised, cloud-native architecture underpinning Pathogenwatch ensures reproducibility, scalability, and operational robustness. Analytical workflows execute consistently across deployments and scale elastically during periods of increased demand. Benchmarking of SARS-CoV-2 lineage assignment demonstrates that these continuously deployed workflows produce results concordant with established standards, supporting their suitability for real-world surveillance.

Pathogenwatch has been adopted as shared analytical infrastructure within multiple genomic surveillance networks. In TyphiNET and KlebNET-GSP (39), Pathogenwatch supports harmonised interpretation of lineage structure and antimicrobial resistance across global collections of *Salmonella enterica* ser. Typhi (6) and *Klebsiella pneumoniae* (7). Within PulseNet Africa, the platform has been integrated into whole-genome sequencing workflows for *Vibrio cholerae*, enabling contextual comparison against global reference data to support food- and water-borne disease surveillance (40). Together, these deployments illustrate how Pathogenwatch facilitates collaboration and analytical consistency across geographically distributed partners while reducing technical barriers to genomic surveillance.

Pathogenwatch’s design and operational practices are consistent with the attributes and principles recently proposed for pathogen genomic data-sharing platforms supporting surveillance of epidemic and pandemic pathogens (41). The platform provides free, unrestricted access through both a graphical interface and a documented RESTful API, with all analytical software publicly available as open-source code. Data provenance is maintained through INSDC accession linking and automated ingestion from international sequence archives, while a privacy and terms-of-service framework operating under the UK Data Protection Act 2018 establishes clear data controller/processor responsibilities. Species-specific quality control derived from QualiBact ensures consistent curation, and the containerised architecture enables rapid incorporation of novel pathogens — positioning Pathogenwatch as infrastructure aligned with emerging international standards for equitable and transparent pathogen genomic data sharing.

## Conclusion

Pathogenwatch provides an accessible, scalable, and reproducible framework for pathogen genome analysis and global genomic surveillance. By combining containerised analytics, continuously curated reference datasets, and a common public health abstraction, the platform transforms raw genomic data into interpretable signals that support surveillance, outbreak investigation, and collaborative analysis across pathogens.

With its transition to a generic, real-time surveillance architecture, Pathogenwatch enables rapid incorporation of pathogens with epidemic or pandemic potential without re-engineering core systems. This positions the platform as durable preparedness infrastructure, capable of supporting both endemic surveillance and rapid genomic response to emerging infectious threats.

## Data Availability

All public data represented in Pathogenwatch are available for download within the application (https://pathogen.watch).

