## supplementary data for "Pathogenwatch: A public health platform for rapid interpretation of pathogen genomics"

### Supplementary methods for: “Pathogenwatch: An always-on genomic surveillance platform for contextualizing pathogen genomes and population structures globally”

Nabil-Fareed Alikhan[1,2], Corin Yeats[1,2], Khalil Abudahab[1,2], Pranit Shinde[1,2], Georgina Lewis-Woodhouse[1,2], Anthony Underwood[1], Silvia Argimón[1], Ravikumar K Lingegowda[3], Pilar Donado-Godoy[4], Sonia Sia[5], Iruka N Okeke[6], Sophia David[1,2], Philip M Ashton[1,2], David M Aanensen[1,2]

**Affiliations:**

1. Centre for Genomic Pathogen Surveillance, Pandemic Sciences Institute, University of Oxford, Oxford, UK
2. WHO Collaborating Centre on Genomic Surveillance of AMR, University of Oxford, Oxford, UK
3. KIMS Hospital and Research Centre, Bangalore, India
4. Global Health Research Unit for the Genomic Surveillance of Antimicrobial Resistance, CI Tibaitatá, Corporación Colombiana de Investigación Agropecuaria (AGROSAVIA), Mosquera, Colombia
5. Research Institute for Tropical Medicine, Manila, Philippines
6. Department of Pharmaceutical Microbiology and Biotechnology, University of Ibadan, Ibadan, Nigeria
7. NIHR Global Health Research Unit on Genomics and enabling data for surveillance of antimicrobial resistance.

#### Speciation with Speciator

Speciator is Pathogenwatch’s in-house tool for assigning a species designation to assembled genomes, which is essential for downstream species-specific analyses (e.g. cgMLST, resistance prediction) (7). It combines a MinHash k-mer search engine (via Mash (8)) with a curated reference library of high-confidence genomes.

When an input genome assembly is submitted, Speciator first searches it against a curated reference library composed of high-confidence genomes drawn from the Kleborate and Bacsort collections (9,10), supplemented with in-house curation. Using a MinHash-based distance metric, the tool compares the input to all entries in this library and assigns a species designation if the closest match falls within a distance threshold of d = 0.04. This initial search provides rapid and accurate classification for most genomes.

If no species-level match meets the threshold, Speciator performs a broader genus-level search against an expanded reference library built from RefSeq genomes. In this step, a more permissive distance threshold (d = 0.15) is applied, and the top twenty matches are retrieved to infer the most likely genus of origin. Once a probable genus is established, the tool refines the classification by re-searching within the corresponding genus-specific reference set, this time using a stricter distance cutoff (d = 0.05) to attempt species-level assignment.

In cases where no confident match can be found, a final search is conducted against a “No Genus” library, again using a distance threshold of d = 0.05. If this final attempt fails to produce a reliable result, the assembly is designated as unclassified. This multi-tiered approach ensures both speed and robustness in species identification, while providing sensible fallbacks for novel or poorly represented taxa.

In tests on thousands of genomes across *Salmonella*, *Klebsiella*, *Neisseria*, and others, Speciator consistently achieved 100% accuracy on curated collections (e.g. SPARK Klebsiella, EuSCAPE non-*K. pneumoniae*, Pathogenwatch public collections). It runs in only a few seconds per assembly, providing rapid, robust input to downstream modules. Speciator is not designed for metagenomic or highly contaminated assemblies (mixed-species genomes will receive a single species label). Moreover, accuracy depends on reference library correctness; misannotated or novel species not yet represented may be misclassified or marked unclassified.
